# Epidemiology of SARS-CoV-2 Emergence Amidst Community-Acquired Respiratory Viruses

**DOI:** 10.1101/2020.07.07.20148163

**Authors:** Karoline Leuzinger, Tim Roloff, Rainer Gosert, Kirstin Sogaard, Klaudia Naegele, Katharina Rentsch, Roland Bingisser, Christian H. Nickel, Hans Pargger, Stefano Bassetti, Julia Bielicki, Nina Khanna, Sarah Tschudin Sutter, Andreas Widmer, Vladimira Hinic, Manuel Battegay, Adrian Egli, Hans H. Hirsch

## Abstract

**Background:** SARS-CoV-2 emerged in China in December 2019 as new cause of severe viral pneumonia (CoVID-19) reaching Europe by late January 2020. We validated the WHO-recommended assay and describe the epidemiology of SARS-CoV-2 and community-acquired respiratory viruses (CARVs).

**Methods:** Naso-oropharyngeal swabs (NOPS) from 7663 individuals were prospectively tested by the Basel-S-gene and the WHO-based E-gene-assay (Roche) using Basel-N-gene-assay for confirmation. CARVs were tested in 2394 NOPS by multiplex-NAT, including 1816 together with SARS-CoV-2.

**Results:** Basel-S-gene and Roche-E-gene-assays were concordant in 7475 cases (97.5%) including 825 (11%) positive samples. In 188 (2.5%) discordant cases, SARS-CoV-2 loads were significantly lower than in concordant positive ones and confirmed in 105 NOPS. Adults were more likely to test positive for SARS-CoV-2, while children were more likely to test CARV-positive. CARV co-infections with SARS-CoV-2 occurred in 1.8%. SARS-CoV-2 replaced other CARVs within 3 weeks reaching 48% of all detected respiratory viruses followed by rhino/enterovirus (13%), influenzavirus (12%), coronavirus (9%), respiratory syncytial (6%) and metapneumovirus (6%).

**Conclusions:** The differential diagnosis for respiratory infections was broad during the early pandemic, affecting infection control and treatment decisions. We discuss the role of pre-existing immunity and competitive CARV replication for the epidemiology of SARS-CoV-2 infection among adults and children.

## Background

Severe acute respiratory syndrome coronavirus-2 (SARS-CoV-2) emerged in China during winter 2019 as new cause of severe viral pneumonia called coronavirus infectious disease (CoVID)-19 [1, 2]. Since late January 2020, SARS-CoV-2 continues to spread across the world including Europe [1, 3]. By the end of May 2020, the WHO reported more than 5 million confirmed SARS-CoV-2 cases, 400 thousand deaths of which approximately one third occurred in Europe [4]. The first case in Switzerland was diagnosed on 25 February 2020, reaching peak rates by the end of March before plateauing at approximately 30’000 confirmed cases by the end of April 2020 (https://covid-19-schweiz.bagapps.ch/de-2.html). Notably, the initial pandemic spread of SARS-CoV-2 occurred in the winter months of the Northern hemisphere, during which several community-acquired respiratory viruses (CARVs) are known to circulate including influenzavirus-A/B, respiratory syncytial virus, metapneumovirus, parainfluenzavirus, and human coronaviruses. Although the progression of SARS-CoV-2 infection to severe lower respiratory tract infectious disease (RTID) is unprecedented, all CARVs are known to significantly contribute to seasonal excess morbidity and mortality in immunocompetent and immunocompromised populations [5]. In fact, CARV-RTID presents clinically as an influenza-like illness defined as at least one respiratory and one systemic symptom/sign such as clogged or runny nasal airways, sore throat, cough, fatigue, fever, myalgia [5], which may be indistinguishable from early stages of SARS-CoV-2 infection [6], while dysgeusia appears to be rather prominent. Thus, a broad diagnostic approach using multiplex nucleic acid testing (NAT) may be important. Notably, the course and impact of SARS-CoV-2 on circulating CARVs has not been fully characterized. Early reports suggested that co-infections with SARS-CoV-2 and other CARVs were rather uncommon in immunocompetent adults [7]. However, a recent study reported co-infection of CARVs and SARS-CoV-2 at rates of 5% including influenzavirus-A/B, respiratory syncytial virus and rhinoviruses [8]. Here, we report on the epidemiology of SARS-CoV-2 infection and other CARVs during the early pandemic peak in Northwestern Switzerland from January 1 until March 29, 2020.

## Methods

### Patients and samples

Patients presenting with influenza-like illness to the outpatient department or emergency department of the University Hospital Basel or the University of Basel Children’s Hospital were enrolled in this analysis of prospectively collected results on respiratory virus panel and/or SARS-CoV-2 testing between 1. January and 29. March 2020.

### Clinical samples and total nucleic acid extraction

For sampling, two swabs from the naso- and oropharyngeal sites (NOPS), respectively, were taken and combined into one universal transport medium tube (UTM, Copan). In smaller children, only nasopharyngeal swabs were taken. Total nucleic acids (TNAs) were extracted from the UTM using the MagNA Pure 96 system and the DNA and viral NA small volume kit (Roche Diagnostics, Rotkreuz, Switzerland) or using the Abbott m2000 Realtime System and the Abbott sample preparation system reagent kit (Abbott, Baar, Switzerland).

### SARS-CoV-2 reverse transcription quantitative nucleic acid testing

TNAs were tested for SARS-CoV-2 RNA using a laboratory-developed reverse transcription quantitative nucleic acid test (RT-QNAT) targeting specific viral sequences of the spike glycoprotein S-gene (Basel-SCoV2-S-111bp; **Supplementary Table 1**) and a commercial RT-QNAT targeting the viral envelope gene (E-gene; Roche). For the Roche assay, the extraction control was included as specified by the manufacturer, while Basel assays were spiked with unrelated nucleic acids. Both assays were run on independent plates in parallel for all NOPS tested. Concordant negative results were interpreted as confirmed, while concordant positive and discordant results were retested with a laboratory-developed RT-QNAT targeting the viral nucleocapsid gene (Basel-SCoV2-N-98bp; **Supplementary Table 1**). The Roche-E-gene was run on the CFX96 RT-PCR (Bio-Rad Laboratories, Cressier, Switzerland) as specified by the manufacturer. The Basel-SCoV2-S-111bp and Basel-SCoV2-N-98bp used the RT Takyon MMX containing uridine and the uracil-N-glycosylase for amplicon decontamination (Eurogentec, Liège, Belgium), 300 nmoles end concentration of the primers and probe for the Basel-S-gene RT-QNAT, and 900 nmoles end concentration of the primers and 100 nmoles probe for the Basel-SCoV2-N-98bp RT-QNAT (**Supplementary Table 1**). The Basel assays had a reaction volume of 25 μL and 5 μL of extracted TNA run on an ABI7500 Fast Real-Time PCR system (Thermo Fisher Scientific, Massachusetts, United States). The single-step reverse transcription and cycling conditions for the RT-QNATs were 50 °C for 10 min; 95 °C for 5 min; and 45 cycles of 95 °C for 30 s and 60°C for 60 s.

**Table 1.** Patients’ demographics of NOPS tested for SARS-CoV-2 between calendar week 11 and 13 (Mann–Whitney U test; n=7663).

|  |  | All | SARS-CoV-2<br>positive | SARS-CoV-2<br>negative | p-value |
| --- | --- | --- | --- | --- | --- |
| Patients' Demographics | Patients (N, %) | 7663 | 930 (12%) | 6733 (88%) | - |
|  | Age (median, IQR) | 43 | 49 | 43 | p<0.001 |
|  |  | 25<sup>th</sup>: 31 | 25<sup>th</sup>: 34 | 25<sup>th</sup>: 30 |  |
|  |  | 75<sup>th</sup>: 58 | 75<sup>th</sup>: 62 | 75<sup>th</sup>: 57 |  |
|  |  | IQR: 27 | IQR: 28 | IQR: 27 |  |
|  | Gender (male, %) | 3407 (44%) | 458 (49%) | 2949 (44%) | P=0.002 |
|  | Pediatric patients<br>(≤16 years) | 354 (5%) | 14 (4%) | 340 (96%) | p<0.001 |
| Primary Care |  | 5697 (74%) | 774 (83%) | 4923 (73%) | - |
| Secondary or Tertiary Care |  | 1966 (26%) | 156 (17%) | 1810 (27%) | - |
|  | Medical Care Unit | 136 (7%) | 11 (7%) | 125 (7%) | - |
|  | Intensive Care Unit | 67 (3%) | 8 (5%) | 59 (3%) | - |
|  | Pneumology Unit | 45 (2%) | 1 (1%) | 44 (2%) | - |

### Phylogenetic analysis of SARS-CoV-2 genome sequences

SARS-CoV-2 complete genome sequences (N=3323), the S-gene and N-gene sequences were downloaded from the NCBI-GenBank and GISAID database (https://www.gisaid.org/; accessed on 20^th^ of April). Sequence alignments were performed with the CLC Genomic Workbench software (version 12; QIAGEN, Hilden, Germany). Divergences were estimated by the Jukes-Cantor method, and neighbor-joining trees were visualized with the CLC Genomic Workbench software. In addition, the frequency of single nucleotide polymorphisms (SNPs) was assessed in the S-gene and N-gene RT-QNAT target regions with the basic variant detection tool of the CLC Genomic Workbench software and a viral reference genome (acc. no. NC_045512; **Supplementary Table 2**) as described previously [9].

**Table 2.** Comparison of SARS-CoV-2 and any CARV infection in adult and pediatric patients using Mann–Whitney U test (n=1816; excluding human rhinovirus only cases n=1671).

| Pathogen | Testresult | Age |  | p-value |
| --- | --- | --- | --- | --- |
|  |  | ≤16 years | >16 years |  |
| SARS-CoV-2 | Positive | 5 (2%) | 143 (9%) | P<0.001 |
|  | Negative | 257 (98%) | 1411 (91%) |  |
| Any CARV<sup>1</sup> | Positive | 166 (63%) | 479 (31%) | P<0.001 |
|  | Negative | 96 (37%) | 1075 (69%) |  |
| SARS-CoV-2 or<br>any CARV | Positive | 169 (65%) | 606 (39%) | P<0.001 |
|  | Negative | 93 (35%) | 948 (61%) |  |
| SARS-CoV-2 or<br>CARV<br>(excluding rhinovirus) | Positive | 127 (65%) | 503 (39%) | P<0.001 |
|  | Negative | 93 (35%) | 948 (61%) |  |

<sup>1</sup>Any CARV includes: human adenoviruses, human coronavirus (-229E, -OC43, -NL63, and -HKU1), influenza virus A and B, human metapneumovirus, human parainfluenzavirus (1 to 4), human rhinovirus, human respiratory syncytial virus and the atypical bacterial agent M. pneumoniae

### Biofire Filmarray respiratory panel

The qualitative multiplex NAT respiratory panel used 200 μL of UTM in the Torch system (Biofire Filmarray respiratory 2.0 panel, bioMérieux) covering influenzavirus (IV)-A (A/H1, A/H1-2009, A/H3), IV-B, human respiratory syncytial virus (HRSV)-A/B, adenovirus (HAdV), human metapneumovirus (HMPV), rhinovirus/enterovirus (HRV), parainfluenzavirus (HPIV) 1-4 (as separate targets), human coronavirus (HCoV)-NL63, -229E, -OC43, -HKU1, -middle east respiratory syndrome (MERS)-CoV, *Bordetella pertussis, B. parapertussis, Chlamydophila pneumonia*, and *Mycoplasma pneumoniae*.

### Statistics and graphical presentations

All statistical data analysis was done in R (https://www.r-project.org/), and Prism (version 8; Graphpad Software, CA, USA) was used for data visualization. Statistical comparison of non-parametric data was done using Mann–Whitney U test, and Bonferroni correction was applied for multiple comparisons.

### Ethics statement

The study was conducted according to good laboratory practice and in accordance with the Declaration of Helsinki and national and institutional standards and was approved by the ethical committee (EKNZ 2020-00769).

## Results

To independently evaluate the WHO-recommended assay, we designed two different single step RT-QNAT assays targeting the S-gene and the N-gene. We observed close clustering of the complete SARS-CoV-2 genomes and specifically its S- and N-gene target sequences, clearly separating from the corresponding HCoV genome sequences (**Supplementary Figure 1**). We found no insertions or deletions in either target, and only a single SNP in the probe-binding site of the S-gene RT-QNAT in one of 3323 (0.03%) sequences, at a central position not predicted to affect the assay performance (**Supplementary Table 2**).

**Figure 1.**
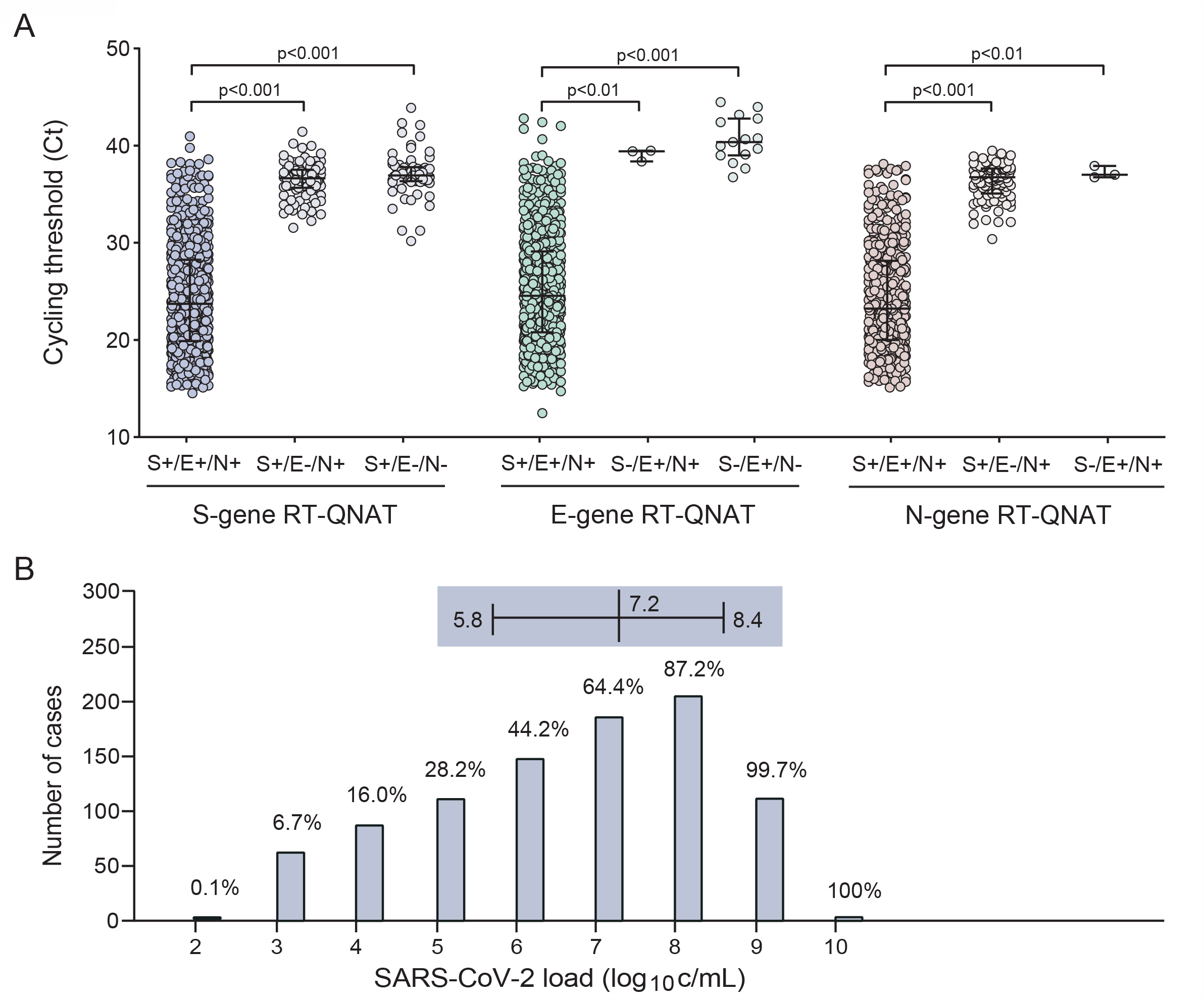
Comparison of cycling thresholds in the S-gene, E-gene and N-gene RT-QNATs and SARS-CoV-2 loads. NOPS were submitted for routine testing with the S-gene and E-gene RT-QNATs (n=7663). Samples with concordant positive or discordant results were subsequently tested with the in-house N-gene RT-QNAT. A. Cycling thresholds of concordant positive and discordant samples are displayed (median, 25^th^ and 75^th^ percentiles; n=7663). B. SARS-CoV-2 loads and number of cases determined with the S-gene RT-QNAT in positive samples (median, 25^th^ and 75^th^ percentiles; n=927).

To cross-validate the WHO-Roche-E-gene and the Basel-S-gene without reporting delay, we analyzed all submitted NOPS directly in parallel. From 9^th^ until 29^th^ of March 2020 (calendar week 11 to 13), 7663 samples were submitted from 354 (5%) pediatric and 7309 (95%) adult patients (**Table 1**). Most patients had presented to primary care and outpatient clinics (74%), while 26% of cases originated from secondary and tertiary care units including 3% from intensive care (**Table 1**). The Basel-S-gene and the Roche-E-gene RT-QNATs were concordant in 7475 (97.5%) samples, consisting of 6650 (86.8%) negative and 825 (10.7%) positive cases, all of which were independently confirmed by the N-gene assay (**Supplementary Figure 2**). In 188 (2.5%) cases, discordant results were obtained consisting of 170 (2.2%) Basel-S-gene positive/ Roche-E-gene negative, and 18 (0.2%) Basel-S-gene negative/Roche-E-gene positive cases. The N-gene RT-QNAT confirmed 102/170 (60%; overall 1.3%) of the former, but only 3 (0.04%) of the latter (**Supplementary Figure 2**). Cycle threshold (Ct)-values were significantly lower in these samples, indicating a higher viral load for concordant positive than for discordant results (median, S-gene RT-QNAT: 23.6 *vs*. 36.8 *vs*. 37.1; P<0.001; **Figure 1A**). Indeed, 666 (72%) NOPS extracts had SARS-CoV-2 loads of more than 1 million copies (c)/mL UTM in the S-gene RT-QNAT (median 7.2 log_10_ c/mL, IQR 5.8 – 8.4; **Figure 1B**). Conversely, the Ct-values were significantly higher for discordant results indicating low viral loads (**Figure 1A**). Thus, the Basel-SCoV2-S-111bp had a sensitivity of 99.7% (95% CI: 95% - 100%) and specificity of 99.0% (95% CI: 91% - 100%). Taken together, 930 (12.1%) SARS-CoV-2 infections were confirmed and further analyzed. Among SARS-CoV-2-positive patients, male gender was more prevalent (49% *vs*. 44%; p=0.002) and the median age was higher (49 *vs*. 43 years; p<0.001) compared to those with a negative test result (**Table 1**). However, higher patient age was not associated with higher SARS-CoV-2 loads (Spearman’s r=0.034, p=0.30). Moreover, SARS-CoV-2 was detected in 14 (4%) of 354 children compared to 916 (12%) of the non-pediatric patients (p<0.05).

**Figure 2.**
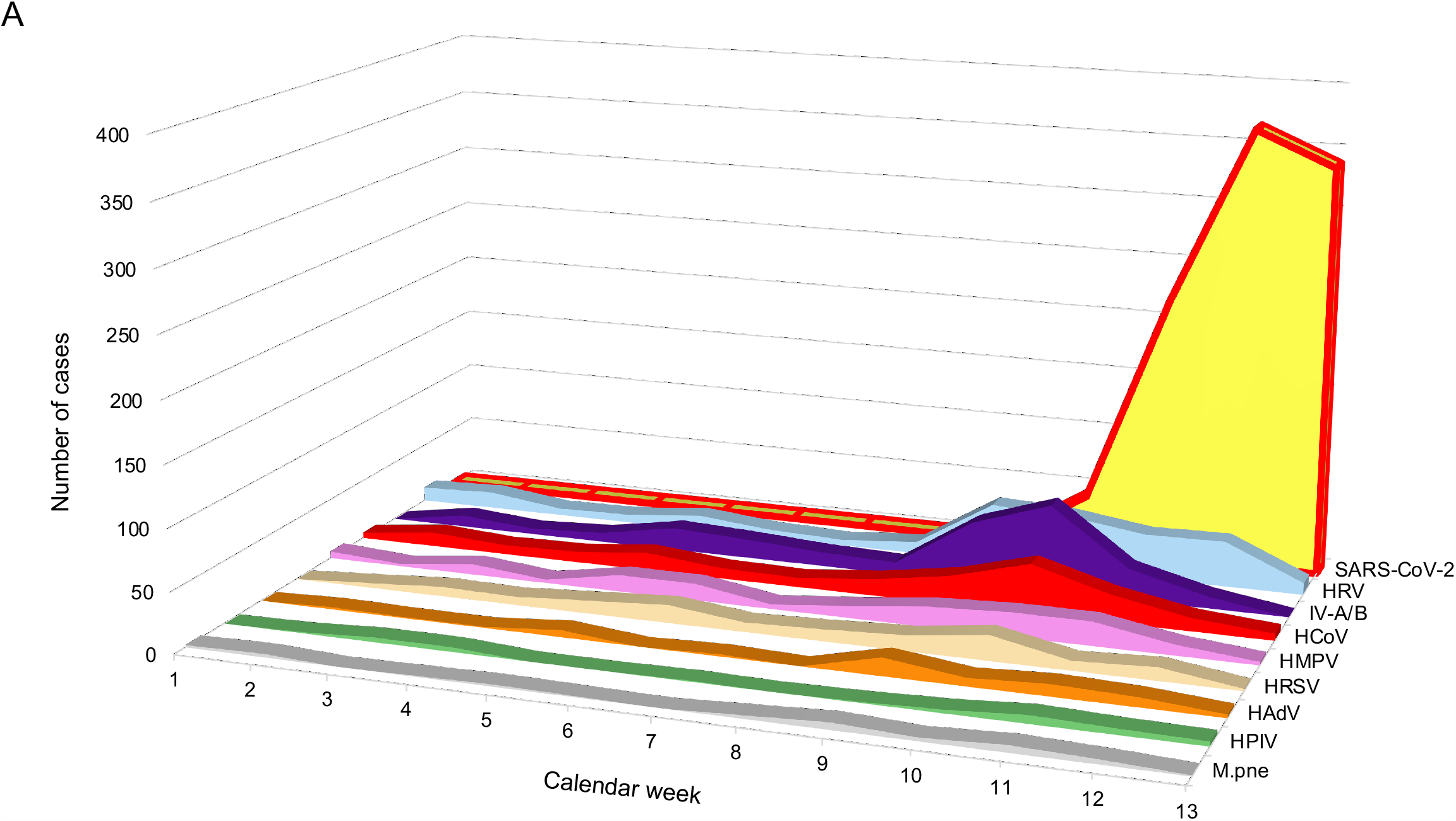
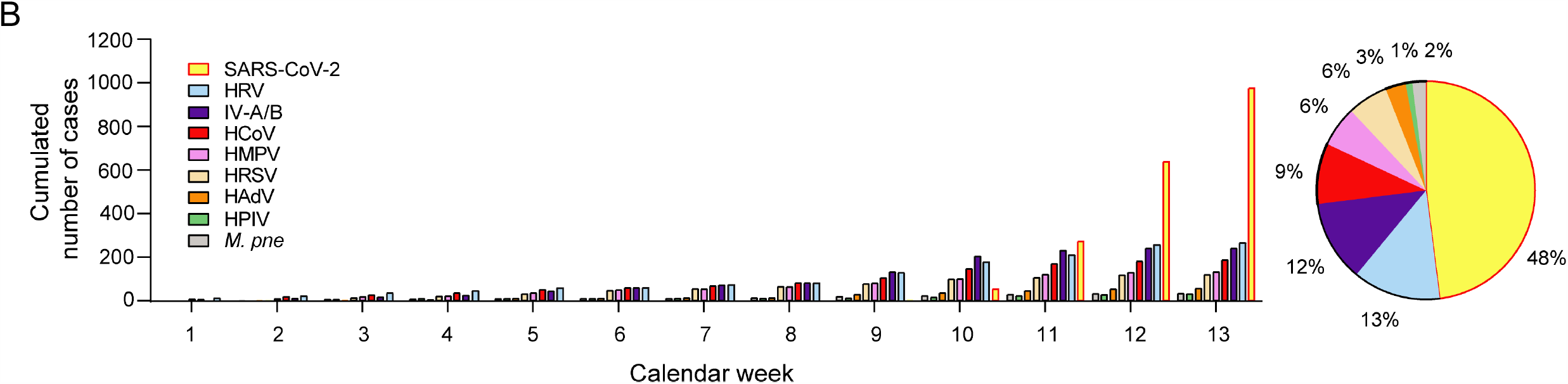
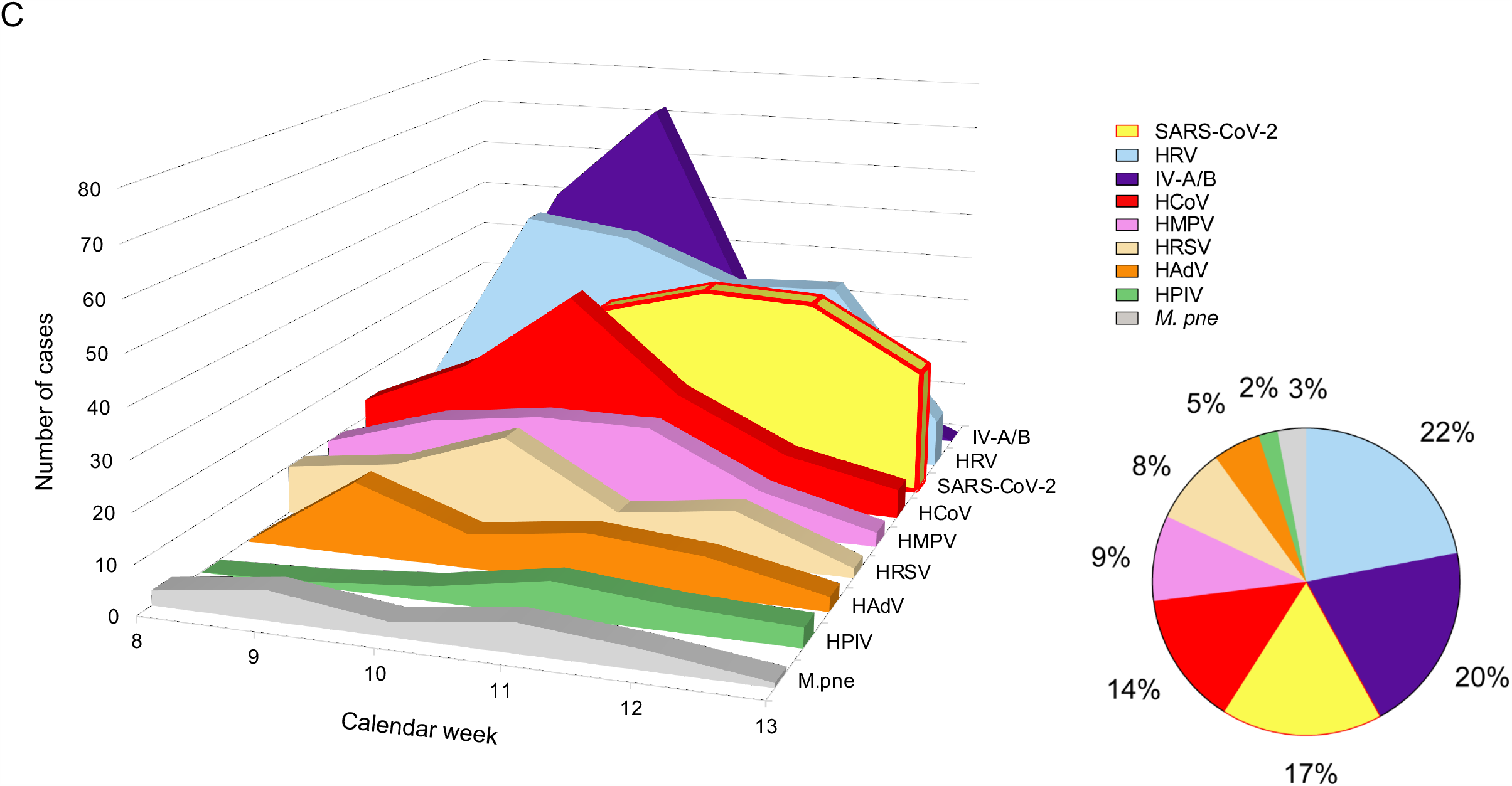
SARS-CoV-2 and CARV epidemiology during the beginning of the epidemic spread from January to March 2020. HAdV, human adenoviruses; HCoV, human coronavirus (−229E, -OC43, -NL63, and -HKU1); IV-A/B, influenza virus A and B; HMPV, human metapneumovirus; HPIV, human parainfluenzavirus (1 to 4); HRV, human rhinovirus; HRSV, human respiratory syncytial virus; M. pne, Mycoplasma pneumoniae; SARS-CoV-2, severe acute respiratory syndrome coronavirus. A. Weekly SARS-CoV-2 (n=8592) and CARV (n=2394) prevalence in symptomatic children and adults. B. Weekly prevalence of cumulated SARS-CoV-2 (n=8592) and CARV (N=2394) cases in symptomatic children and adults (bar chart), cumulated SARS-CoV-2 and CARV cases by calendar week 13 (pie chart). Weekly SARS-CoV-2 and CARV prevalence in NOPS tested in parallel (n=1816), cumulated SARS-CoV-2 and CARV cases by calendar week 13 (pie chart).

To investigate the epidemiology of CARVs and SARS-CoV-2 during the first phase of the pandemic, we identified all NOPS (n=2394) from patients with influenza-like illness, which had been tested by CARV-multiplex-NAT between January 1^st^ (calendar week 1) and March 29^th^ 2020 (calendar week 13). In 942 (39%) cases, at least one pathogen had been detected including 95 with two (3.9%), and 9 with 3 (0.1%) pathogens. The weekly prevalence rates for SARS-CoV-2 and CARVs revealed a fluctuating CARV activity until calendar week 7 followed by a steep increase in CARVs, which declined after week 10, when SARS-CoV-2 detection rates rose sharply (**Figure 2A**). This was also reflected in the cumulative rates (**Figure 2B; histogram**) reaching 48% for SARS-CoV-2 by calendar week 13, followed by rhinovirus (13%) and influenzavirus (12%) (**Figure 2B; pie chart**). Restricting the analysis to 1816 NOPS, from which both, CARV-multiplex-NAT and SARS-CoV-2 RT-QNAT had been requested, the cumulative SARS-CoV-2 detection was 17% after rhinovirus (22%) and influenzavirus (20%) (**Figure 2C; pie chart**). The weekly detection rates revealed that SARS-CoV-2 largely replaced all other CARVs except rhinovirus (**Figure 2C**).

Unlike for SARS-CoV-2, the CARV detection rate was significantly higher in children than in adults (P<0.001; **Table 2**). This significant effect also prevailed when SARS-CoV-2 and CARV positive cases where analyzed together and when excluding rhinovirus-infected cases from the analysis (**Table 2; see also below**). Analyzing the age distribution of CARV-positive cases (**Figure 3**), we found higher detection rates of adenovirus, parainfluenzavirus, respiratory syncytial virus, rhinovirus and influenzavirus-A/B cases in children ≤16 years, while similar rates of human coronavirus-, metapneumovirus- and *M. pneumoniae* were detected in children and adults (**Supplementary Table 3**). Moreover, adenovirus-positive patients were significantly younger than patients testing positive for other CARVs or SARS-CoV-2 (P<0.001; **Table 3**). Among adults (1554/1816; 85.5%), no significant age differences were observed for patients testing positive for any CARV, but patients being positive for SARS-CoV-2 tended to be older than patients testing positive for any other CARV (P<0.01; **Figure 3, Supplementary Table 4**).

**Table 3.**
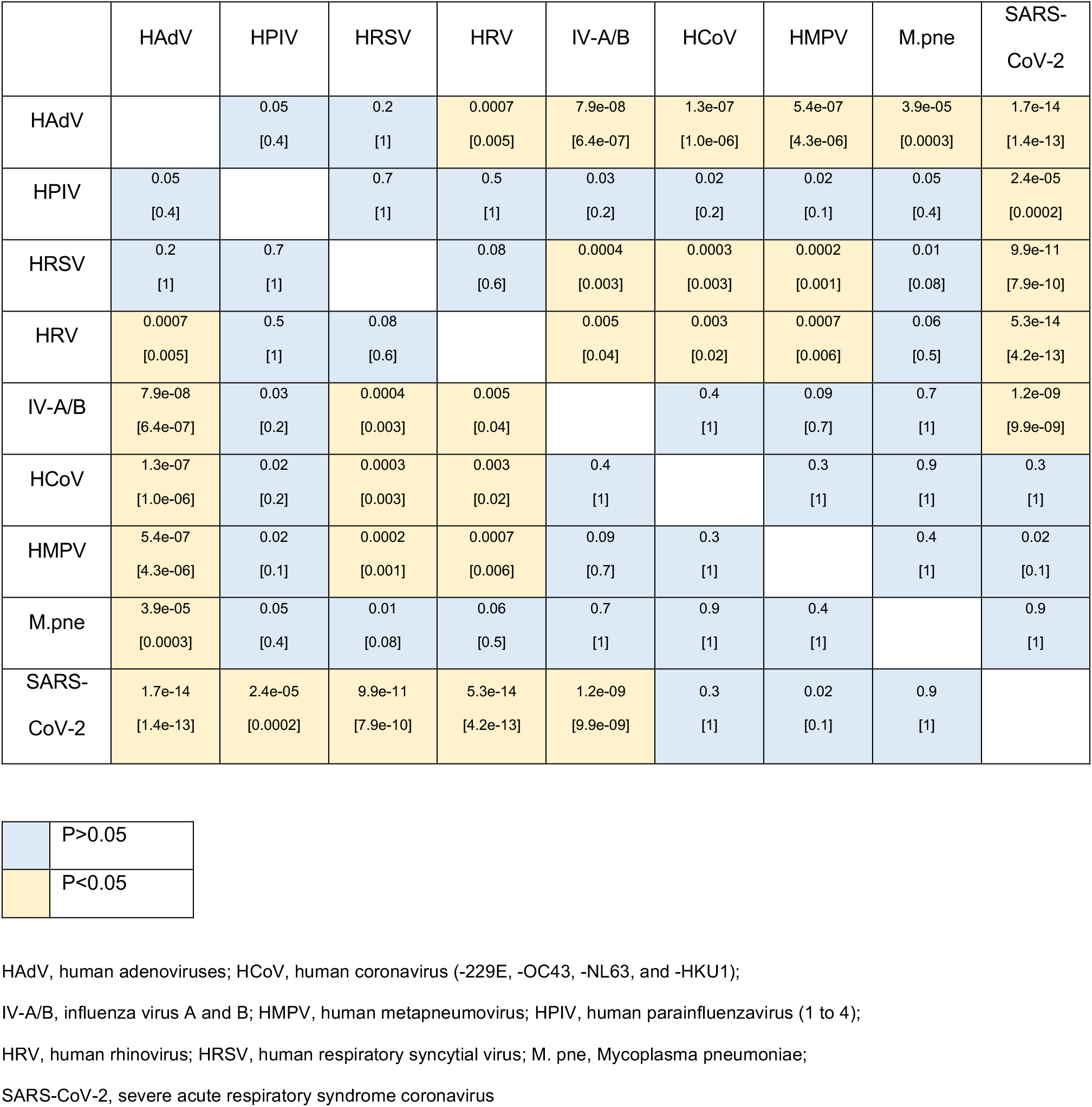
Pairwise comparison of patient age in SARS-CoV-2 or CARV positive patients (Mann–Whitney U test; brackets indicate the p-value after Bonferroni correction; n=1816).

**Table 4.**
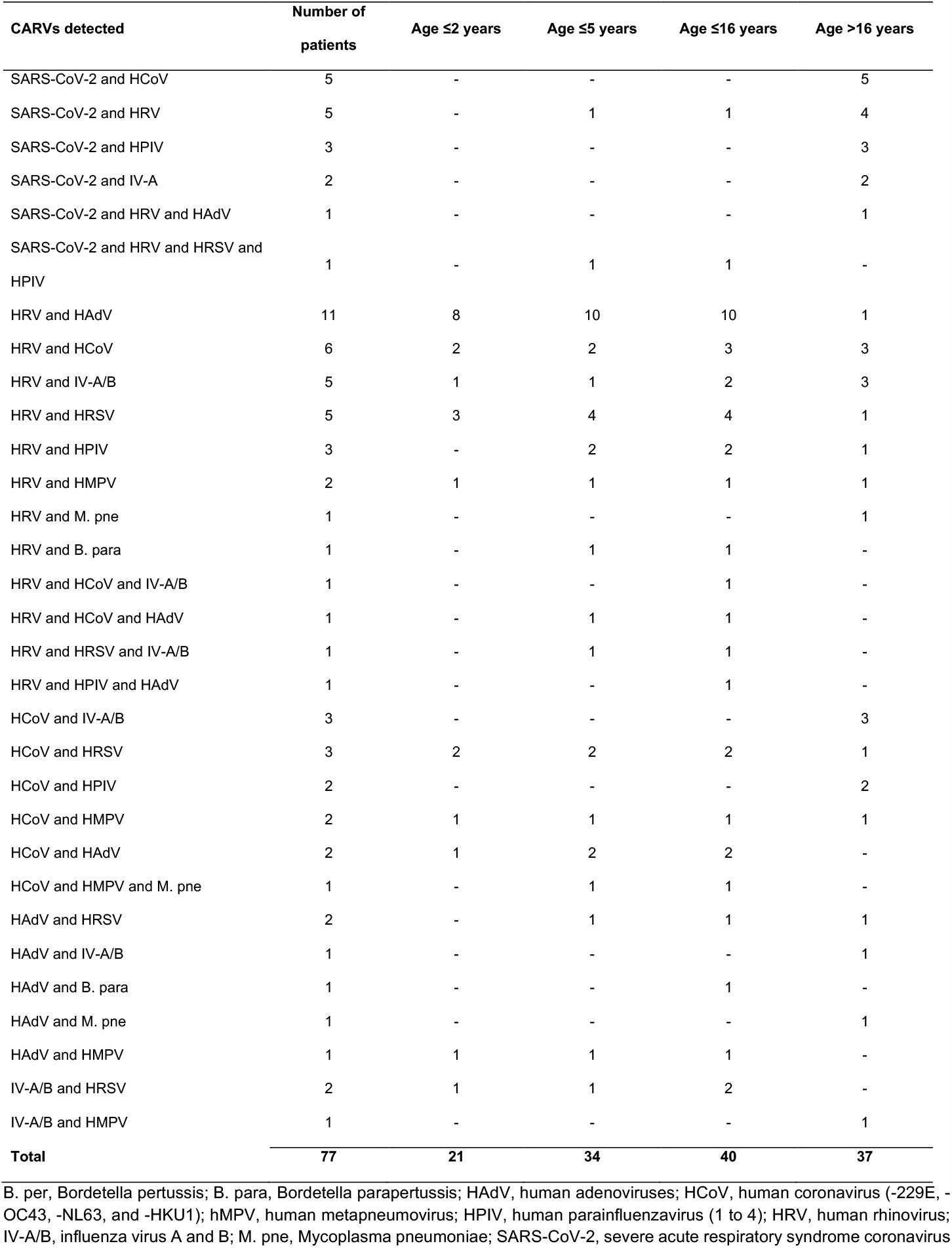
Patients with more than one positive SARS-CoV-2 or CARV detection (n=1816).

| CARVs detected | Number of patients | Age ≤2 years | Age ≤5 years | Age ≤16 years | Age >16 years |
| --- | --- | --- | --- | --- | --- |
| SARS-CoV-2 and HCoV | 5 | - | - | - | 5 |
| SARS-CoV-2 and HRV | 5 | - | 1 | 1 | 4 |
| SARS-CoV-2 and HPIV | 3 | - | - | - | 3 |
| SARS-CoV-2 and IV-A | 2 | - | - | - | 2 |
| SARS-CoV-2 and HRV and HAdV | 1 | - | - | - | 1 |
| SARS-CoV-2 and HRV and HRSV and HPIV | 1 | - | 1 | 1 | - |
| HRV and HAdV | 11 | 8 | 10 | 10 | 1 |
| HRV and HCoV | 6 | 2 | 2 | 3 | 3 |
| HRV and IV-A/B | 5 | 1 | 1 | 2 | 3 |
| HRV and HRSV | 5 | 3 | 4 | 4 | 1 |
| HRV and HPIV | 3 | - | 2 | 2 | 1 |
| HRV and HMPV | 2 | 1 | 1 | 1 | 1 |
| HRV and M. pne | 1 | - | - | - | 1 |
| HRV and B. para | 1 | - | 1 | 1 | - |
| HRV and HCoV and IV-A/B | 1 | - | - | 1 | - |
| HRV and HCoV and HAdV | 1 | - | 1 | 1 | - |
| HRV and HRSV and IV-A/B | 1 | - | 1 | 1 | - |
| HRV and HPIV and HAdV | 1 | - | - | 1 | - |
| HCoV and IV-A/B | 3 | - | - | - | 3 |
| HCoV and HRSV | 3 | 2 | 2 | 2 | 1 |
| HCoV and HPIV | 2 | - | - | - | 2 |
| HCoV and HMPV | 2 | 1 | 1 | 1 | 1 |
| HCoV and HAdV | 2 | 1 | 2 | 2 | - |
| HCoV and HMPV and M. pne | 1 | - | 1 | 1 | - |
| HAdV and HRSV | 2 | - | 1 | 1 | 1 |
| HAdV and IV-A/B | 1 | - | - | - | 1 |
| HAdV and B. para | 1 | - | - | 1 | - |
| HAdV and M. pne | 1 | - | - | - | 1 |
| HAdV and HMPV | 1 | 1 | 1 | 1 | - |
| IV-A/B and HRSV | 2 | 1 | 1 | 2 | - |
| IV-A/B and HMPV | 1 | - | - | - | 1 |
| **Total** | **77** | **21** | **34** | **40** | **37** |

B. per, Bordetella pertussis; B. para, Bordetella parapertussis; HAdV, human adenoviruses; HCoV, human coronavirus (-229E, -OC43, -NL63, and -HKU1); hMPV, human metapneumovirus; HPIV, human parainfluenzavirus (1 to 4); HRV, human rhinovirus; IV-A/B, influenza virus A and B; M. pne, Mycoplasma pneumoniae; SARS-CoV-2, severe acute respiratory syndrome coronavirus

**Figure 3.**
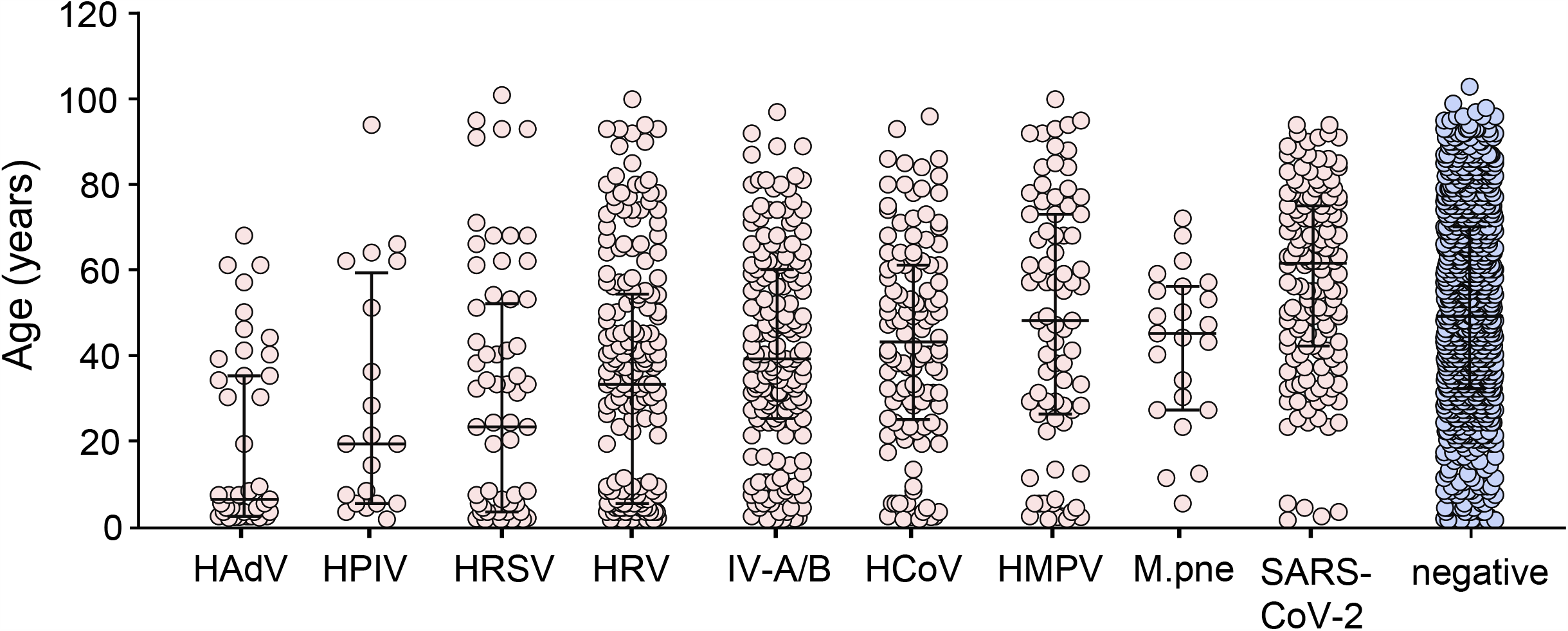
Age distribution of CARV and SARS-CoV-2 positive patients. NOPS were analyzed in parallel for different CARVs with multiplex NAT and SARS-CoV-2 with the RT-QNAT assays. Patient age of CARV or SARS-CoV-2 positive patients is displayed (median, 25^th^ and 75^th^ percentiles; n=1816), and compared using Mann–Whitney U test (Table 3; Supplementary Table 4).

Among CARV positive cases, co-infections with two or three CARVs occurred in 55 (3%) and five (0.3%) patients, respectively (**Table 4**). Rhinovirus and adenovirus as well as rhinovirus and respiratory syncytial virus co-infections were almost exclusively found in children ≤2 years. In 17 (0.9%) of 1816 patients (15 adults aged 30 to 93 years, 2 children), SARS-CoV-2 was detected together with at least one other CARV, which consisted of a single pathogen in 15 cases, namely rhinovirus (n=5), human coronaviruses (n=5), parainfluenzavirus (n=3) and influenzavirus (n=2), and more than one CARV detection in 2 cases (**Table 4**). Overall, CARV detection was associated with a high negative predictive value of 98.1% for SARS-CoV-2 infection. The negative predictive value for SARS-CoV-2 infection was higher in CARV-positive children (99.0%; ≤16 years) than adults (97.1%; >16 years). Conversely, a negative multiplex-NAT result after the first detected SARS-CoV-2 case was associated with a rapidly increasing likelihood to be positively tested for SARS-CoV-2 from 1% in calendar week 9 to 48% in calendar week 13 (**Figure 2**).

## Discussion

The SARS-CoV-2 pandemic hit Europe in winter 2020, during which a number of circulating CARVs are at their yearly seasonal peak including influenzavirus, respiratory syncytial virus, and human coronaviruses. Our analysis from the start to the peak of the pandemic wave of SARS-CoV-2 in Northwestern Switzerland has three major findings.

First, the early pandemic phase until calendar week 10 was dominated by winter CARVs, emphasizing the importance of their rapid and accurate identification due to several reasons: i) significant morbidity and mortality in vulnerable patients (very young, elderly, immunocompromised) [5]; ii) specific antiviral therapy in case of influenzavirus-A/B detection; iii) appropriate infection control and cohorting strategies upon hospital admission; and iv) prevention of unnecessary empiric antibiotic therapy in CARV-positive patients, or treatment adaptation in case of atypical bacterial agents like *M. pneumoniae* [10–13]. In this early phase, CARV detection was associated with a high negative predictive value of 98.1% for SARS-CoV-2 infection.

Second, SARS-CoV-2 almost completely replaced the seasonally circulating CARVs within only 3 weeks’ time. During calendar week 12 and 13, SARS-CoV-2 was practically the only respiratory virus leading to a cumulative 48% runner-up position when counting all detected CARVs from January 1^st^, 2020. This dynamic evolution was also seen when explicitly analyzing NOPS from patients with respiratory illness for whom both CARV- and SARS-CoV-2 testing had been requested. The weekly prevalence revealed a significant increase in SARS-CoV-2 detection rates while the initially increasing CARVs were curtailed. These data suggest the intriguing possibility of competing risks for host infection.

Third, diagnosis of SARS-CoV-2 infection was highly reliable being based on three independent molecular tests. Thereby, an independent validation of the WHO-endorsed E-gene was provided. Whereas respiratory panel testing is well validated and widely used in tertiary care centers [5, 14, 15], the response to the SARS-CoV-2 pandemic hinges on the performance of a new diagnostic test for a new viral agent, and its communication within a short turn-around time. To accomplish this task, we prospectively tested all NOPS directly in parallel with the commercial Roche-E-gene and our Basel-S-gene RT-QNAT. This outstanding opportunity for independent test validation on more than 7600 patients demonstrated high concordance of 97.5% between both assays including 825 (11%) SARS-CoV-2 infections, which could be communicated without further delay to the treating physicians. Importantly, comparison of the Ct-values revealed that the discordance mostly resulted from SARS-CoV-2 loads at the limit of detection. Thus, discordant results became increasingly likely at very low, hence limiting viral loads in the NOPS, most likely reflecting a stochastic distribution of genomes in the analyte.

A limitation of our study is the dependence on the pre-analytic steps of NOPS sampling, especially in the light of the natural course of SARS-CoV-2 infection. We addressed this challenge through repeated instructions and video clips demonstrating the correct use of personal protection equipment, validated swab sets, and defined sampling procedures in dedicated hospital areas. However, late presentation at stages of more advanced disease manifesting in the lower respiratory tract requires testing of respiratory samples from the lower respiratory tract, while this study was restricted to the first diagnostic testing in NOPS. Early testing is clinically and epidemiologically advisable in view of high viral loads detectable already early in the course in exposed pre-symptomatic and in oligo-symptomatic persons [16, 17].

Since our diagnostic laboratory is serving both regional tertiary care centers for adults and children, we examined the age distribution of SARS-CoV-2 and CARV infection. Indeed, patients testing positive for adenovirus and respiratory syncytial virus were significantly younger and more likely to be children below the age of 5 years, among whom SARS-CoV-2 infection remained rare, in line with other studies [18, 19]. Although SARS-CoV-2 positive adults were older than patients testing positive for CARVs, the median age of >40 years in CARV-positive patients suggests that similar adult populations were at risk for established CARVs or for the novel SARS-CoV-2.

Our study also provides intriguing observations regarding the epidemiology of SARS-CoV-2 in its capacity to replace circulating CARVs among adults. Notably, co-infection rates of CARVs with SARS-CoV-2 were rather low as reported here and by others [8], suggesting a competitive infection situation. It is presently unclear whether virus properties such as higher infectiousness, facilitated transmission, or increased host susceptibility are the decisive factors conferring significant advantages to the novel SARS-CoV-2 in this first wave of the pandemic. Regarding the infectiousness of SARS-CoV-2, our data provide independent evidence for very high viral loads in the order of 1 – 100 million copies per milliliter transport medium. Even if these high numbers only carry 1000-fold lower infectious units, the infectious activity remains high in the patients’ respiratory secretions. Notably, similarly high viral loads have also been described for CARVs including influenza or respiratory syncytial virus [5, 20–22]. Regarding transmission, SARS-CoV-2 is thought to behave less like influenzaviruses spreading significantly by aerosols [23, 24], but rather like respiratory syncytial virus spreading by droplets, contaminated surfaces and hands [25, 26]. However, aerosolization of SARS-CoV-2 may also play a role, especially when associated with high-velocity air streams during sneezing, singing and medical procedures [27–29]. Finally, increased susceptibility of the human host to infection by this novel, presumably zoonotic coronavirus remains, but is a difficult to estimate factor at this time. Already the first reports from China in January 2020 indicated that SARS-CoV-2 is well adapted to the human host [30]. Unlike SARS-CoV-1, SARS-CoV-2 is easily transmitted from human to human already before the start of symptoms, hence facilitating the pandemic spread [17, 31, 32]. However, SARS-CoV-2 seems to be susceptible to type-1 interferons [33], and induces high amounts of interleukin-6 through excessive macrophage activation upon progression to viral pneumonia, which has become a clinically relevant target of CoVID-19 treatment [34].

What could be the underlying mechanisms for an increased susceptibility to SARS-CoV-2 infection competitively replacing established CARVs in a mostly adult population? Hand washing, social distancing and lock-down measure would be predicted to affect CARVs and SARS-CoV-2 alike, but were not yet sufficiently effective to prevent the upswing of the pandemic wave. We hypothesize that the decisive factors may be the differential net response of the host to virus-induced unspecific innate immunity on the one hand and to virus-specific adaptive immune memory on the other hand. CARV infections are known to cause an innate immune response including type-1 interferons, which reduces the risk of co-infection by other viruses including SARS-CoV-2 [35, 36]. Since adults have been repeatedly exposed to CARVs in the past, their CARV-specific immune memory may not be high enough to prevent symptomatic CARV re-infection, but is readily boosted upon re-exposure, hence limiting CARV replication and the associated inflammation elicited by innate immunity. We propose that thereby the semi-immune mostly adult host population becomes available for SARS-CoV-2 infection. Since SARS-CoV-2 is novel having little, if any, specific immune memory, its replication is prolonged, evoking pronounced inflammation, delaying infection by other circulating CARVs, extending transmission periods and shifting the epidemiologic curve in favor of this novel agent. This differential net response of virus-induced unspecific innate immunity and virus-specific adaptive immune memory may also contribute to the puzzling lower infection rates seen in small children, who typically replicate CARVs in high frequency and high levels, hence interfering with SARS-CoV-2. However, CARVs may differ in their propensity to interfere and may be low for rhinovirus. Indeed, 46 (60%) of 77 co-infections involved rhinovirus. Although other (co-)factors cannot be excluded, our hypothesis will be testable by analyzing, whether or not vaccines to CARVs and/or to SARS-CoV-2 change this competitive epidemiologic risk. Possibly, CARV interference will be reduced during the summer months putting younger age populations at risk for the pandemic SARS-CoV-2.

In conclusion, circulating CARVs were dominant during the first phase of the CoVID-19 pandemic, but rapidly replaced within 2 weeks by SARS-CoV-2. A comprehensive testing strategy covering SARS-CoV-2 and CARVs is central to infection control and clinical management. Epidemiologic determinants of the competitive infection risk between established CARVs and the novel SARS-CoV-2 require further studies including unspecific innate immune interference and virus-specific adaptive immune memory.

## Data Availability

all data referred to in the manuscript are available

## Acknowledgements

We thank the biomedical technicians of the Clinical Virology Division and the Clinical Bacteriology and Mycology Division, Laboratory Medicine, University Hospital Basel, Basel, Switzerland for expert help and assistance.

---

**Figure S1.**
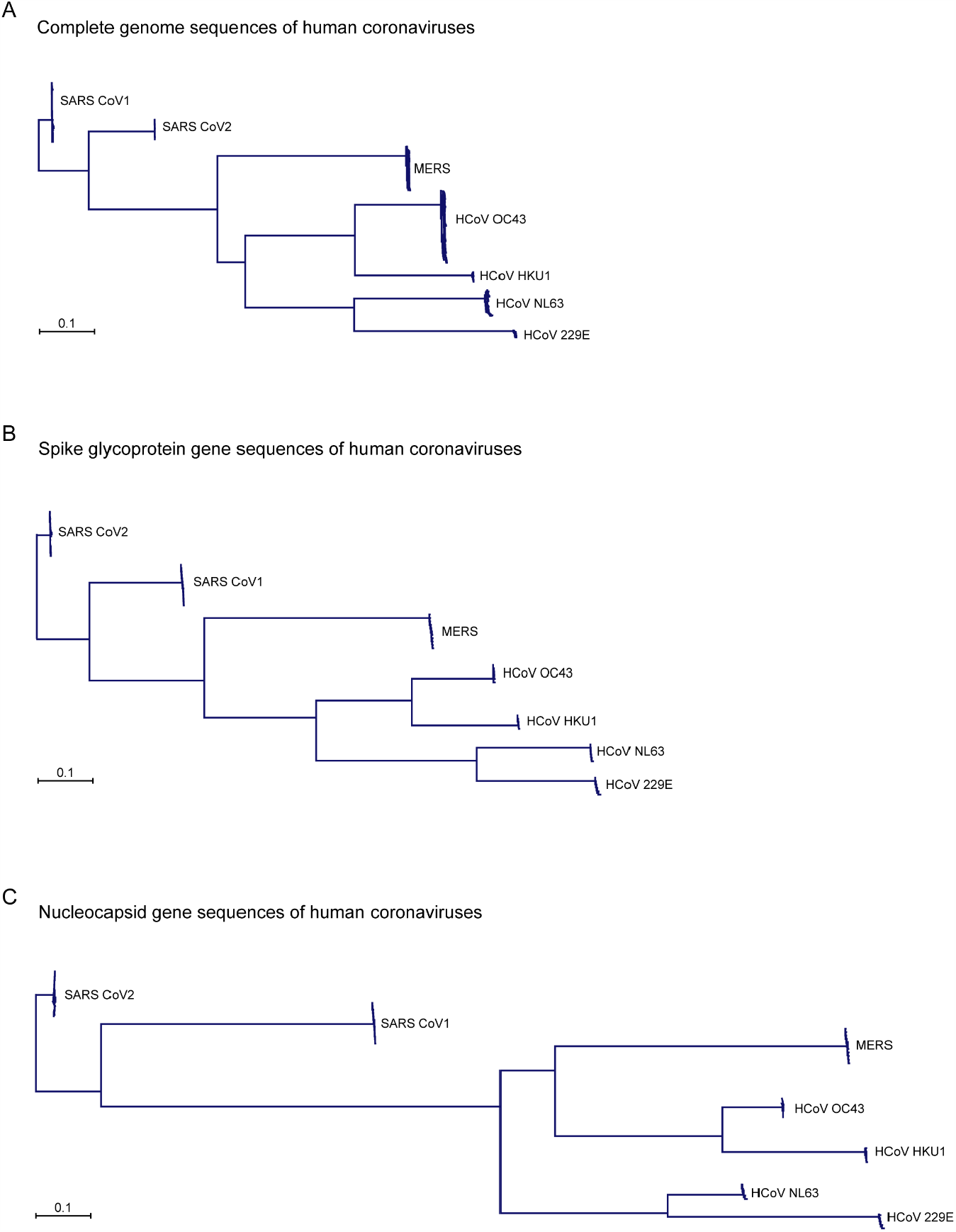
Phylogenetic analysis using complete SARS-CoV-2 genome sequences, the *S-*gene and N-gene sequences. Divergences were estimated by the Jukes-Cantor method and neighbor-joining trees were constructed with the CLC Genomic Workbench software. A. Phylogenetic analysis of complete SARS-CoV-2 genome sequences of 29’900 nucleotides (nt) in length available in the NCBI-GenBank and GISAID database (accessed on 20^th^ of April; n=3323). B. Phylogenetic analysis of the S-gene region of 3’822 nt (corresponds to nucleotide positions 21563 to 25384 in the SARS-CoV-2 reference genome [acc. no. NC_045512.2]; n=3323). C. Phylogenetic analysis of the N-gene region of 366 nt (corresponds to nucleotide positions 27894 to 28259 in the SARS-CoV-2 reference genome [acc. no. NC_045512.2]; n=3323).

**Figure S2.**
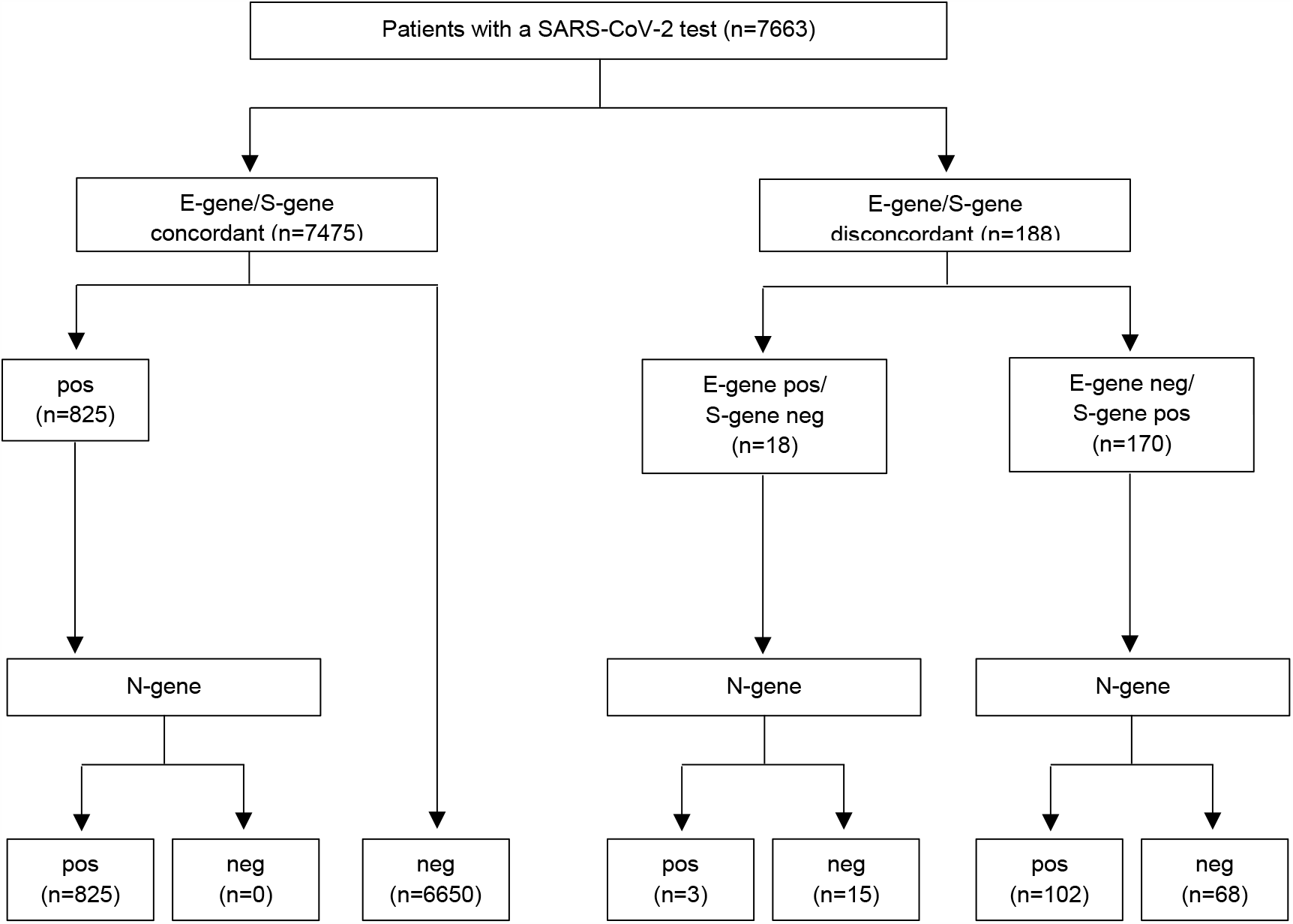
SARS-CoV-2 testing flowchart. NOPS were tested in parallel with the in-house S-gene and the commercial E-gene RT-QNATs (n=7663). Samples with concordant positive or discordant results were subsequently tested with the in-house N-gene RT-QNAT. pos, positive; neg, negative.

**Table S1.**
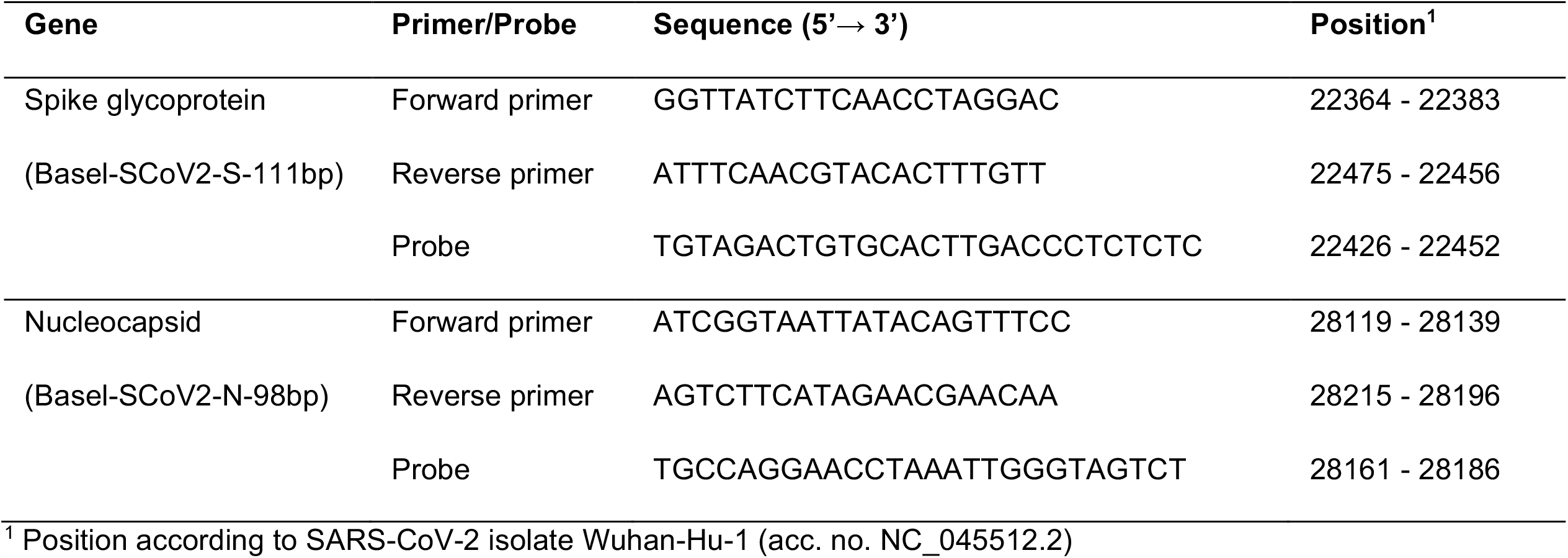
Forward primers, reverse primers and probes of the SARS-CoV-2 RT-QNATs.

**Table S2.** Frequency of single nucleotide polymorphisms in the laboratory-developed SARS-CoV-2 spike glycoprotein (Basel-SCoV2-S-111bp) and nucleocapsid (Basel-SCoV2-N-98bp) RT-QNAT target regions.

| Gene | Position<sup>1</sup> | SNP<sup>2</sup> | Frequency (%) |  |  |
| --- | --- | --- | --- | --- | --- |
|  |  |  | ±150bp<sup>3</sup> | ±100bp<sup>3</sup> | ±5bp<sup>3</sup> |
| Spike glycoprotein<br>(Basel-SCoV2-S-111bp) | 22224 | C → G | 0.03%<sup>4</sup> | - | - |
|  | 22303 | T → G | 0.03%<sup>4</sup> | 0.03%<sup>4</sup> | - |
|  | 22432 | C → T | 0.03%<sup>5</sup> | 0.03%<sup>5</sup> | 0.03%<sup>5</sup> |
| Nucleocapsid<br>(Basel-SCoV2-S-98bp) | 28077 | G → C | 0.1%<sup>4</sup> | 0.1%<sup>4</sup> | - |
|  | 28144 | T → C | 0.3%<sup>6</sup> | 0.3%<sup>6</sup> | 0.3%<sup>6</sup> |

<sup>1</sup> Position according to SARS-CoV-2 isolate Wuhan-Hu-1 (acc. no. NC\_045512.2)
<sup>2</sup> Single nucleotide polymorphism (SNP) and frequency among 3323 SARS-CoV-2 (taxid: 2697049) spike glycoprotein-gene and nucleocapsid-gene GenBank and GISAID sequences (accessed on 20<sup>th</sup> of April 2020) compared to SARS-CoV-2 as reference genome (acc. no. NC\_045512.2).
<sup>3</sup> RT-QNAT target region ±150bp, ±100bp and ±5bp
<sup>4</sup> SNP outside the RT-QNAT target region
<sup>5</sup> SNP in the probe-binding site
<sup>6</sup> SNP in the RT-QNAT target region, but not in a primer/probe-binding site

**Table S3.** Comparison of SARS-CoV-2 and any CARV infection and the age distribution of adult and pediatric patients using Mann–Whitney U test (n=1816).

| Pathogen | ≤16 years<br>(N=262) |  | >16 years<br>(N=1554) |  | p-value |
| --- | --- | --- | --- | --- | --- |
|  | Number of positive<br>cases | Age | Number of positive<br>cases | Age |  |
|  |  | Distribution |  | Distribution |  |
| HAdV | 27 (10%) | Median: 2 | 16 (1%) | Median: 40 | P<0.001 |
|  |  | 25<sup>th</sup>: 1 |  | 25<sup>th</sup>: 34 |  |
|  |  | 75<sup>th</sup>: 4 |  | 75<sup>th</sup>: 51 |  |
|  |  | IQR: 3 |  | IQR: 17 |  |
| HPIV | 9 (3%) | Median: 4 | 11 (1%) | Median: 50 | P=0.02 |
|  |  | 25<sup>th</sup>: 3 |  | 25<sup>th</sup>: 24 |  |
|  |  | 75<sup>th</sup>: 6 |  | 75<sup>th</sup>: 62 |  |
|  |  | IQR: 3 |  | IQR: 39 |  |
| HRSV | 29 (11%) | Median: 1 | 36 (2%) | Median: 42 | P<0.001 |
|  |  | 25<sup>th</sup>: 1 |  | 25<sup>th</sup>: 32 |  |
|  |  | 75<sup>th</sup>: 4 |  | 75<sup>th</sup>: 66 |  |
|  |  | IQR: 3 |  | IQR: 34 |  |
| HRV | 71 (27%) | Median: 3 | 119 (8%) | Median: 44 | P<0.001 |
|  |  | 25<sup>th</sup>: 1.5 |  | 25<sup>th</sup>: 36 |  |
|  |  | 75<sup>th</sup>: 4.5 |  | 75<sup>th</sup>: 67 |  |
|  |  | IQR: 3.0 |  | IQR: 31 |  |
| IV-A/B | 34 (13%) | Median: 6 | 135 (9%) | Median: 46 | P=0.03 |
|  |  | 25<sup>th</sup>: 3 |  | 25<sup>th</sup>: 32 |  |
|  |  | 75<sup>th</sup>: 9 |  | 75<sup>th</sup>: 62 |  |
|  |  | IQR: 6 |  | IQR: 30 |  |
| HCoV | 18 (7%) | Median: 3 | 100 (6%) | Median: 49 | P=0.79 |
|  |  | 25<sup>th</sup>: 1 |  | 25<sup>th</sup>: 34 |  |
|  |  | 75<sup>th</sup>: 4 |  | 75<sup>th</sup>: 63 |  |
|  |  | IQR: 3 |  | IQR: 29 |  |
| HMPV | 16 (6%) | Median: 4 | 63 (4%) | Median: 58 | P=0.13 |
|  |  | 25<sup>th</sup>: 1 |  | 25<sup>th</sup>: 38 |  |
|  |  | 75<sup>th</sup>: 4 |  | 75<sup>th</sup>: 76 |  |
|  |  | IQR: 3 |  | IQR: 38 |  |
| M. pne | 3 (1%) | Median: 10 | 20 (1%) | Median: 47 | P=0.85 |
|  |  | 25<sup>th</sup>: 7 |  | 25<sup>th</sup>: 38 |  |
|  |  | 75<sup>th</sup>: 11 |  | 75<sup>th</sup>: 55 |  |
|  |  | IQR: 4 |  | IQR: 18 |  |
| SARS-CoV-2 | 5 (2%) | Median: 2 | 143 (9%) | Median: 61 | P<0.001 |
|  |  | 25<sup>th</sup>: 1 |  | 25<sup>th</sup>: 43 |  |
|  |  | 75<sup>th</sup>: 3 |  | 75<sup>th</sup>: 75 |  |
|  |  | IQR: 2 |  | IQR: 32 |  |

HAdV, human adenoviruses; HCoV, human coronavirus (-229E, -OC43, -NL63, and -HKU1);
IV-A/B, influenza virus A and B; HMPV, human metapneumovirus; HPIV, human parainfluenzavirus (1 to 4);
HRV, human rhinovirus; HRSV, human respiratory syncytial virus; M. pne, Mycoplasma pneumoniae;
SARS-CoV-2, severe acute respiratory syndrome coronavirus

**Table S4.**
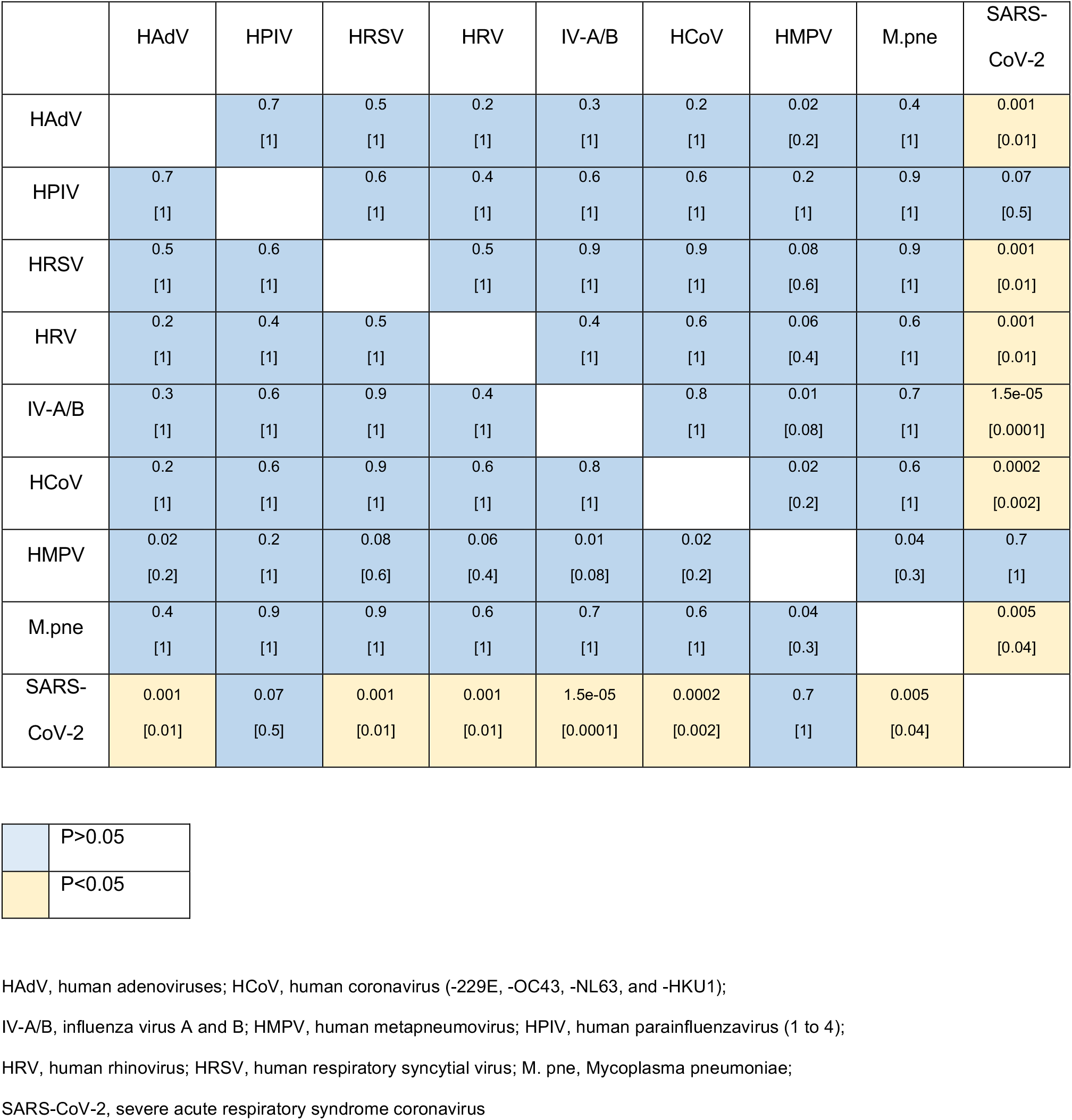
Pairwise comparison of patient age in SARS-CoV-2 or CARV positive adults of >16 years (Mann–Whitney U test; brackets indicate the p-value after Bonferroni correction; n=1554).

